# COVID-19: making the best out of a forced transition to online medical teaching. A mixed methods study

**DOI:** 10.1101/2021.01.19.21249790

**Authors:** Montserrat Virumbrales, Marta Elorduy, Mariona Graell, Pau Mezquita, Pedro Brotons, Albert Balaguer

## Abstract

**Introduction:** the COVID-19 pandemic resulted in a decreed confinement in our country from March until the end of term in June 2020. This forced a transition exclusively to distance learning. The aim of this study was to broaden the understanding of fully online distance learning from the experiences of undergraduate medical students and faculty members during confinement, and identify its key elements.

**Methods:** A convergent mixed methods study analyzing: (a) an online teaching follow-up program, (b) two focus groups and a nominal group with students and faculty, respectively, and (c) a survey with students from 1st to 5th year.

**Results:** Thirteen strongly interconnected categories were identified. Four played an organizational role: course planning, coordination, communication and pedagogical coherence. The remaining nine categories were: learning outcomes, teaching methodology, online resources, evaluation, time management, workload, student motivation, participation, and teacher-student relationship. Among the key aspects of learning were those that promoted rapport between faculty and students, such as synchronous sessions, especially those based on clinical cases.

**Conclusion:** the experiences from confinement allowed us to gain insight into some of the key aspects of online medical teaching. Promoting student motivation and participation at all levels was essential to distance learning in Medicine.

## Introduction

The novel Coronavirus-19 infectious disease (COVID-19) pandemic has dramatically changed the economic, social and daily life of millions of people worldwide since it emerged in Wuhan, China, in December 2019 [1]. One of the countries most affected by COVID-19 has been Spain, with over 778,000 cases and 31,900 deaths confirmed as of October 1, 2020 [2]. A state of emergency was declared by the national government on March 14, imposing strict confinement for the population and closure of most businesses and all leisure, cultural and educational places across the country. As the wave of the pandemic slowly began to recede, confinement was gradually eased until the complete lifting of lockdown restrictions on June 21.

The disruption of learning activities has particularly impacted graduate medical education, an academic field where practical skill acquisition and collaborative training experiences become central [3-5]. While clinical clerkships constitute the primary learning format in the final phase of the undergraduate medical curriculum, face-to-face modalities are essential to acquire basic knowledge and competences at initial stages. As a consequence of the critical situation caused by the pandemic, a number of programmed clinical clerkships had to be urgently readjusted so that senior undergraduates could give support to the clinical management of COVID-19 at a diversity of primary care centers and hospitals. In contrast, lockdown of schools of Medicine resulted in face-to-face learning becoming suddenly halted at initial undergraduate courses. In addition, university faculty, most of which were also deeply involved in the healthcare provision to COVID-19 patients, were committed to transforming face-to-face learning to a virtual format with little or no previous experience in distance learning environments. Boards of medical schools across the nation faced compelling challenges to replace or combine in-person learning with online formats while addressing students’ needs and expectations, in anticipation of future scenarios of potential COVID-19 re-emergence [6]. However, little is known about the effects of such measures.

Upon the inception of confinement, the Universitat Internacional de Catalunya (UIC) School of Medicine (Barcelona, Spain) established an online teaching follow-up program to anticipate the needs of both faculty and students and to ensure that the teaching objectives were met. Through this program, the Dean’s Office continuously monitored the evolving situation and made decisions as changes were required. Out of the total amount of teaching activities traditionally carried out in classrooms (including laboratories and simulation rooms), approximately 60% took place using live online platforms, whereas the remaining 40% was provided in pre-recorded format. The use of virtual classroom activities was encouraged at all levels, particularly those that promoted active student involvement such as games and apps, pop quizzes, online tutoring of individual or group projects, etc. An effort was made to ensure that students had the technical means to access the teaching materials on the virtual learning platform, while making sure that these materials contained the necessary resources adapted to the specific situation (short videos, audio embedded presentations, external resources, etc.). In addition, communication channels were tightened between students, student delegates (2 per year), faculty and the Dean’s Office regarding methodology, resources, student workload and evaluation criteria.

In the midst of this process, we were interested in finding ways to analyze the outcomes of such deep and abrupt changes in the teaching and learning experience. Identifying the lessons learned from this unplanned teaching revolution became crucial to better prepare faculty members for future scenarios in which full on-campus medical education may not be feasible. It also constituted a great opportunity to incorporate the best online practices as complements to face-to-face education once a scenario of normality could be recovered.

Taken all this into account we set out to carry a study with two objectives: (a) to describe and analyze the perceptions and experiences of faculty members and undergraduate medical students (1^st^ to 5^th^ year) as a consequence of switching from on-campus to full online education following the country-wide COVID-19 confinement, and (b) to identify the key elements that need to be addressed in successful online medical learning.

## Materials and methods

A single-center prospective study was conducted with undergraduate students and faculty members at UIC School of Medicine during the months of June and July of 2020. The study site is an academic institution with a strong commitment to educational innovation and extensive adoption of technology-based learning methods, located in the metropolitan area of Barcelona. High COVID-19 incidence rates have been reported in this area since February 2020.

The school enrolls 100 new medical students every year. It offers a fully integrated medical curriculum from 1st to 5th year that includes clinical rotations (3^rd^ to 5^th^ year) and a final clerkship in the sixth year, which was excluded from this study.The study was approved by the Ethics and Research Committee of the study site and written informed consents were obtained from all participants prior to initiation.

The study was conducted through 3 phases using a convergent mixed-methods approach [7,8]:

### Phase 1: Analysis of the data gathered by the online teaching follow-up program (OTFP)

The OTFP program was established at the beginning of the confinement to anticipate the needs of both faculty and students during this period and to ensure that the teaching objectives were met. Through this program, three members of the Dean’s office (DO) continuously monitored the evolving situation and made the appropriate decisions as required. The program relied on the monitoring of a set of key elementsgathered from: a) periodic meetings held separately with faculty, student delegates and representatives of student organizations; b) a cloud-based registry shared between the DO, subject coordinators and student representatives containing reports of incidents and the progress for each subject, and c) a periodic meeting between the DO and a group of student representatives of all courses to make adjustments to methodology, workload and evaluation criteria.

The categorization of the core aspects identified, along with the review of the relevant academic literature, constituted the basis of the student online survey and the script used in the focus group and nominal group techniques conducted with students and faculty respectively in phase 2.

### Phase 2

A) Two focus group discussions were carried out with 22 purposely chosen medical students from first to fifth year: one group of first- and second-year medical students, and another one with third-, fourth- and fifth-year medical students. On another hand, one nominal group discussion was held with 8 faculty members with diverse fields of expertise, technological skills and levels of academic performance.

Criteria for selection of participants in focus and nominal groups sought to obtain the highest quantity and quality of the information provided, as well as avoid any potential age and gender bias. The discussion group online meetings were moderated by two medical faculty and health education researchers, and recorded and transcribed with the acquiescence of all the participants.

B) Anonymous online survey conducted with students from 1^st^ to 5^th^ year of Medicine (n=483) who had received on-campus education until confinement. Prior to its launch, the survey was reviewed by the team of researchers for comprehension and refinement of wording. In addition to questions about demographics, the survey used five-point Likert-scaled ratings and open-ended questions about activities used in online teaching, motivators, expectations, prioritizations, recommendations, positive and negative aspects of the methodology employed, communication methods, good and bad practices, and a global opinion on the teaching and learning experience [9].

### Phase 3: Processing and triangulation of qualitative and quantitative data

The qualitative data of the focus and nominal groups were analyzed using the Atlas-ti program 8.4.4. Distance learning experiences lived by students and faculty were interpreted from the perspective of their protagonists [10]. The data were summarized and classified within the previous key elements allowing for the emergence of categories and subcategories, and labelled accordingly.

Quantitative responses to the online survey were summarized as means and standard deviations (scoring variables) and proportions (nominal variables). The significance of differences between scores of quantitative responses by course or age group was assessed using the Student t-test or the ANOVA test. Statistical significance was set at a p-value of < 0.05. All quantitative analyses were performed using Stata v.15.1 software (Stata Corp., USA).

Data analysis and interpretation was carried out independently by three researchers and later shared through discussion to validate the information that encompasses the different dimensions of the learning transformation, integrate the data with those obtained from other sources and validate the consistency of the qualitative findings. Exploitation of these data aimed to identify the key elements for online learning.

## Results

### Phase 1

The analysis of the data gathered by the OTFP, along with the review of the literature, allowed the identification of the following key teaching aspects: a. student motivation; b. pedagogical coherence; c. teaching methodology; d. online resources; e. student participation; f. teacher-student relationship; g. workload; h. learning outcomes.

### Phase 2a

A total of 22 students participated in the focal groups, which were predominantly composed by women (63.6%) with a uniform representation of students from all classes: 1^st^ yr. (n=4), 2^nd^ yr. (n=5), 3^rd^ yr. (n=5), 4^th^ yr. (n=4) and 5^th^ yr. (n=4). The group was representative of the full spectrum of students in terms of academic performance, participation and collaboration with the DO. The nominal group involved 8 faculty members representing different fields of knowledge from 1^st^ to 5^th^ year that had participated in distance teaching during confinement. Their ages ranged from 40 to 62 years, with different levels of experience and a men: women ratio of 3:1, similar to the proportion found among teachers in the degree.

The analysis carried out in the focal and nominal group talks allowed a categorization of the data where, in addition to the 8 categories identified in phase 1, which served as the starting point, 5 new categories emerged: course planning, coordination, communication, time management, and evaluation. A few elements complemented pre-existing categories rather than created new ones. These elements were classified as “subcategories” and are described in table 1, along with their corresponding descriptors and number of quotations.

**Table 1.**
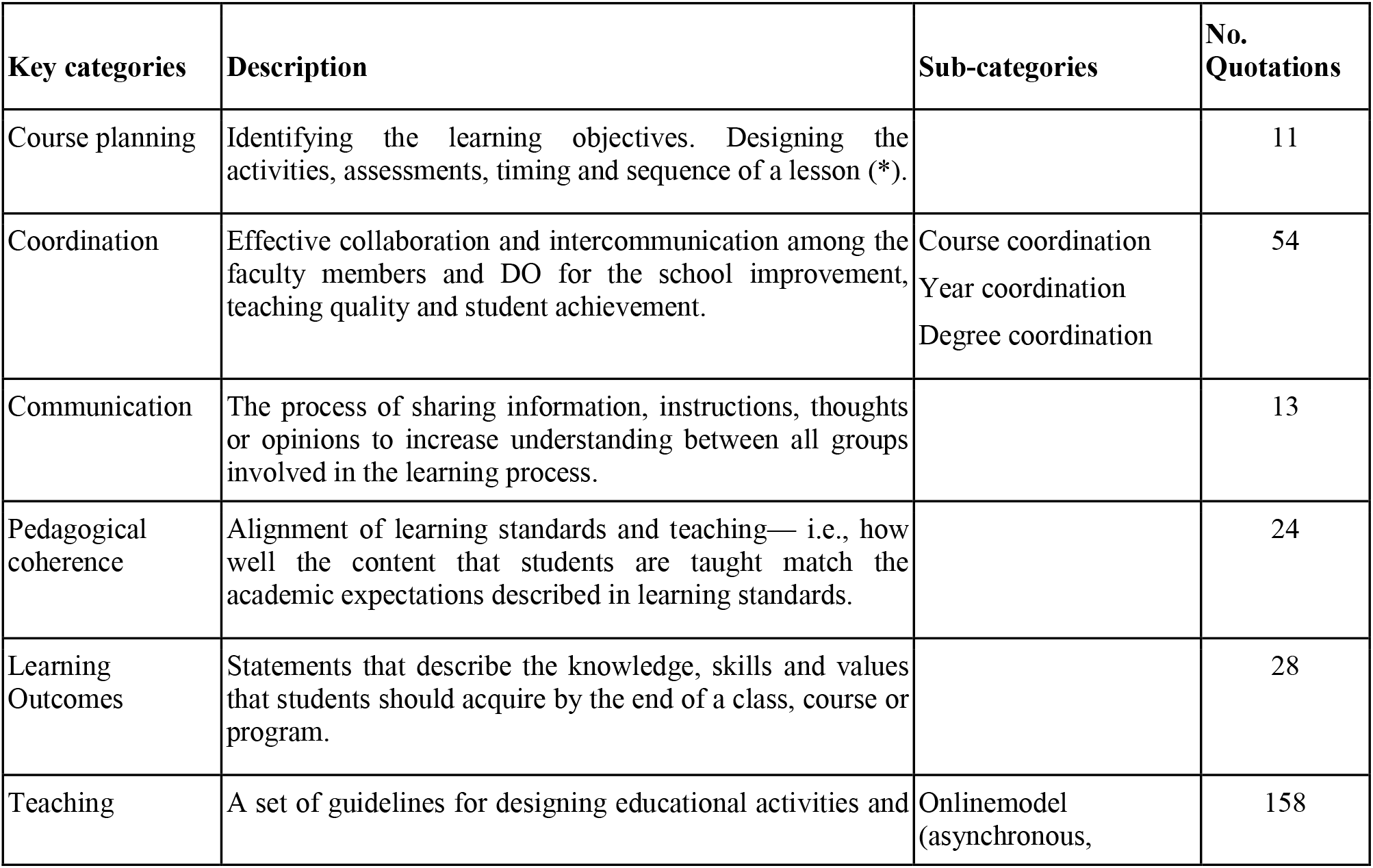
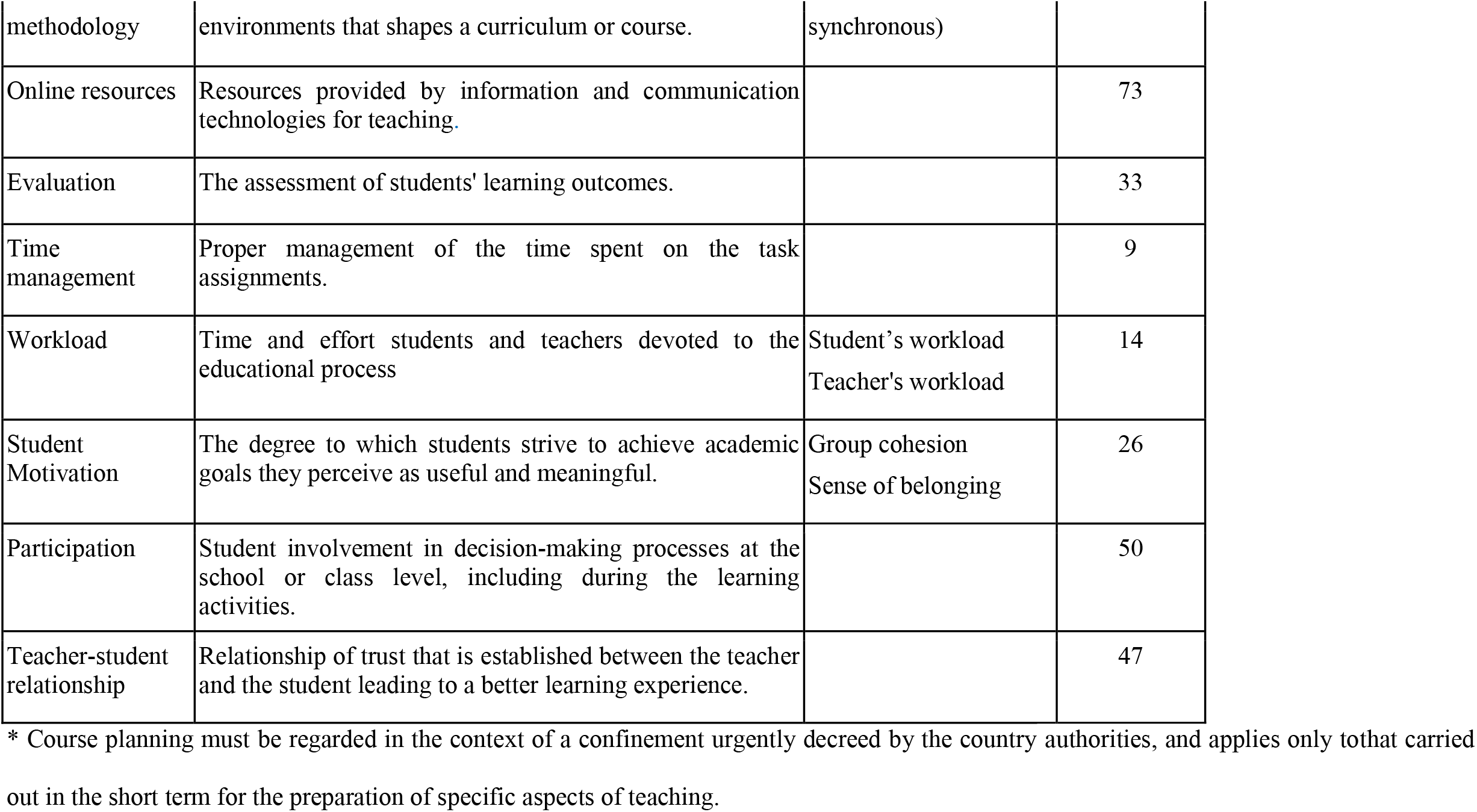
Categories, descriptors, subcategories and number of quotations obtained from qualitative analysis of discussion groups with faculty and students

These thirteen categories have helped us to understand the perception of teaching during this critical period from the perspective of both students and teachers, providing us with an insight on the limitations and difficulties encountered as well as the positive aspects, both described in table 2.

**Table 2.** Main perceptions of students and faculty about online versus traditional face-to-face teaching.

| Aspects | Students | Teachers |
| --- | --- | --- |
| Positive/<br>Strengths | ▪ greater learning autonomy<br>▪ better use of time<br>▪ better conciliation with social life<br>▪ learning at your own pace (through teacher-supplied resources, such as recorded classes, supplementary notes, etc.)<br>▪ direct contact in synchronous sessions | ▪ work-life conciliation (e.g. fewer trips)<br>▪ increased diversity in ways of communicating with students<br>▪ encourages reflection and focusing in the most important aspects |
| Difficulties/<br>Limitations | ▪ remaining motivated<br>▪ participating actively<br>▪ preserving the student-teacher relationship<br>▪ understanding the pedagogical coherence of new assignments<br>▪ excessive workload<br>▪ frequent improvisation in teaching.<br>▪ changes in the calendar/non-compliances<br>▪ loss of social life in campus | ▪ shift in the role of teaching.<br>▪ loneliness (i.e. during online sessions)<br>▪ increased workload<br>▪ obtaining feedback from students<br>▪ adapting to the dynamics of synchronous online sessions (multitasking)<br>▪ lack of digital skills<br>▪ student personal knowledge (needs, achievements, ...)<br>▪ adapting materials in a short period of time |

During phase 2b, which was carried out simultaneously with the focal and nominal groups, we conducted a survey of students with a 50.5% response rate (n=244). On a Likert scale of 1 to 5 (ranging from “worst” to “best”), students from 1st and 2nd years rated distance learning as acceptable (average value of µ=3.1; SD-1.2), while those in 3rd, 4th and 5th years rated it as unsatisfactory (µ=2.7; SD-1.1); (p<0.01). A total of 63.1% of students stated that they had “learned less” with online teaching compared to face-to-face interaction. The worst rated aspect was the additional student workload involved in distance learning (µ=0.7; SD=0.7). Some of other poorly valued aspects were: teacher-student relationship (µ=2.1; SD=1.1), motivation (µ=2.3; SD=0.1), planning (µ=2.6; SD=1.0), pedagogical coherence (µ=2.6; SD=1.0) and participation (µ=2.9; SD=1.3). The aspects that received the highest scores for optimized learning were lectures carried out using synchronic communication tools (µ=3.9; SD=1.3) and case-based teaching (µ=3.4; SD=1.2).

Comparison of the results obtained from both quantitative and qualitative methods allowed us to identify and qualify a number of key aspects that may contribute to an optimal teaching and learning experience, which are discussed in the following section. Figure 1 details the interconnections between the categories identified.

**Figure 1.**
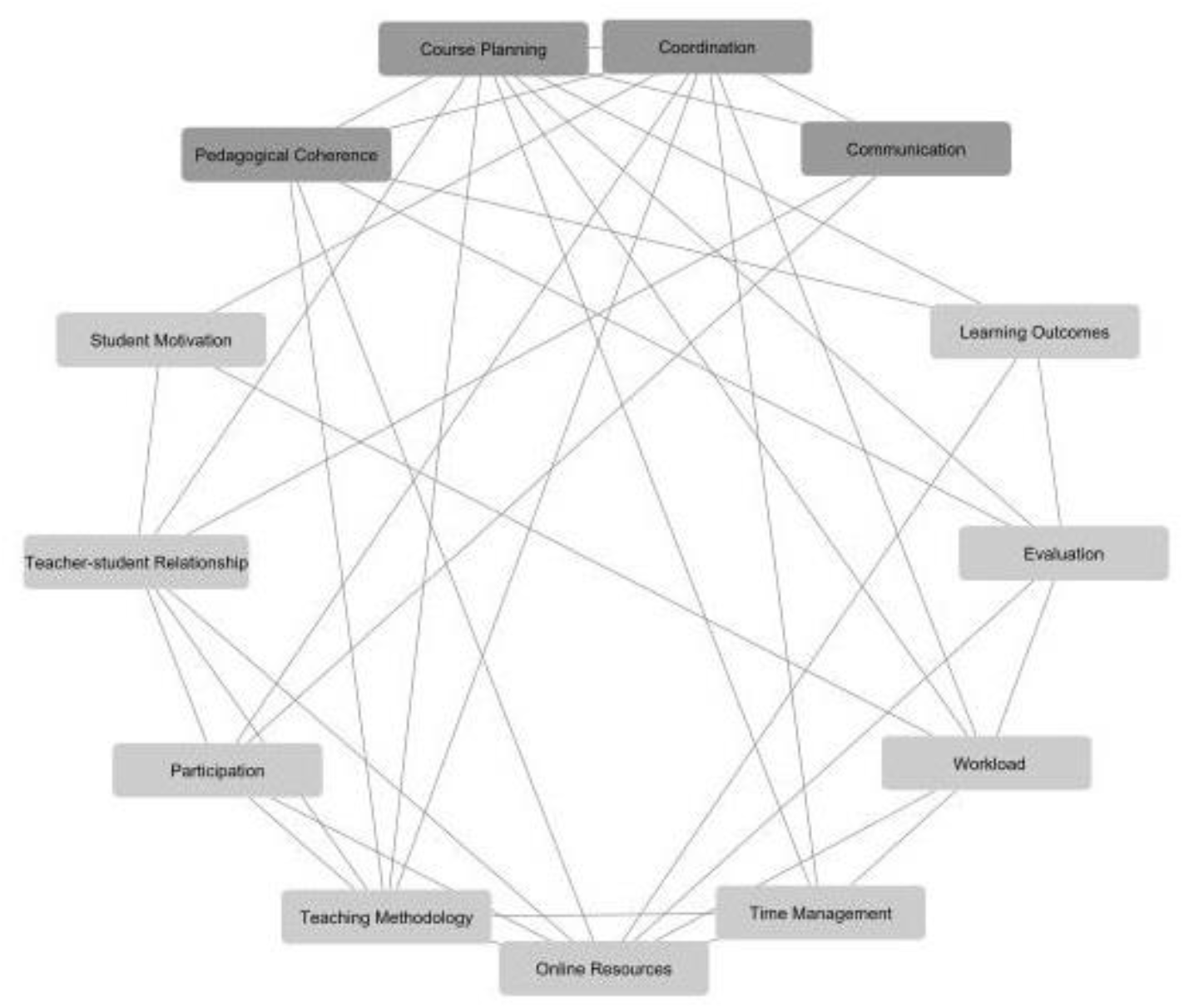
Relationship between categories.

## Discussion

Overall, most categories identified through our phased analysis were aligned with core themes of online medical learning described in the pre-COVID-19 era [11]. We speculate that the educational disruption caused by the pandemic probably aggravated negative aspects traditionally associated with distance learning and undermined those that could be more beneficial for students and faculty. Perceptions and experiences of students and faculty in relation to each category are discussed in the following subsections.

### Course planning

This category emerged as a key element among all discussion groups when implementing any change in the teaching process. The sudden shift from face-to-face to distance learning made the adoption of quick and clear guidelines to face the new situation a daunting task. This resulted in the spread of a feeling of uncertainty among students regarding schedules, teaching methodology, assigned tasks and evaluation criteria. Students expected faculty to provide upfront information and answers on how the course would proceed in the light of the developing events:

> *“*… *I felt a constant uncertainty of not knowing the deadlines well enough, nor the weight of the many tasks that were assigned to us. The same applied to some exams*…*” (student)*
>
> *“*… *we had to build our own schedule on the go, because there was really none set ahead of time. Sometimes we were given a one-day notice on a lecture, or even on the same day” (student)*

The survey corroborated the importance of planning and its low rating among students (2.6 out of 5). The deficits in course planning and their immediate consequences highlight the need to provide contingency plans shared by students and faculty to face similar situations in the foreseeable future.

As far as the faculty is concerned, the experience was lived as an opportunity to reflect on teaching, its means and its purpose. A “positive” counterpoint amid the dramatic situation:

> *“Despite all the misfortune, from a teacher’s perspective this has become an opportunity. It has forced us to question many things we took for granted. [It has given us] a chance to rethink aspects of teaching that were considered already settled” (faculty)*

### Coordination

The school had in place a well-defined coordination structure prior to the crisis, which included classrepresentatives, subject coordinators (some subjects involved 6 or more faculty members), year coordinators and the DO. The fact that a large portion of faculty were attending physicians caught in a major health crisis only added to the challenge of having to make right decisions and carry out proper actions, as well as providing the students with fast and reliable information on these actions. Despite the efforts, many students described a feeling of restlessness concerning their academic lives, which was aggravated by the emotional toll the crisis was taking on the students themselves, their relatives and the society in general.

> *“*… *the role played by some subject coordinators was poor… I believe they could have shown more involvement, [they could have] acted more like subject coordinators… and communicated better with us, through the class representatives or by any other means”* (*student)*
>
> *“*… *it was a very very stressful process for everyone to go through because, besides the confinement and everything that was happening, [we had] fears, doubts…” (student)*

These feelings of stress were described by some students despite the preventive measures and psychological support provided by the university to students and faculty, in line with other reports [12,13].

### Communication

In the midst of such an unprecedented crisis, online teaching required more than ever an effective and multi-directional communication strategy that would allow the spread of information between all the parts involved (students, faculty, the DO and staff). Many of the challenges experienced could have been alleviated or avoided by establishing a good level of communication.

> *“Communication, especially in the beginning, was a large area for improvement*… *particularly among faculty and between faculty and students. There were many [communication] channels and information [provided by these channels] was often misaligned*…. *there were quite a fewmisunderstandings*…*” (student)*

Outside of the synchronous online classes, the most commonly used method of communication between faculty and students was e-mail. This method proved to have its own limitations. In addition, communication was held to a large extent with class representatives exclusively, which left many students feeling that communication was insufficient and wishing they had received information first hand. In contrast, initiatives held by some faculty that took advantage of alternative methods of communication such as synchronous chats or asynchronous forums were highly praised.

### Pedagogical coherence

This aspect was highlighted above all by students, many of whom expressed the difficulty in relating some of the activities proposed to the corresponding learning objectives. Consequently, pedagogical coherence scored low in the survey (2.6/5). A similar perception was observed regarding the weight or impact of certain tasks in the evaluation process.

> *“*… *it was very important for me to understand the importance of many activities that were assigned to us, as well as how they weighed in with regards to the final subject examination. This made things very difficult for me in terms of time management” (student)”*

### Learning Outcomes

Faculty and students both expressed the subjective feeling that they accomplished less through distance learning than in face-to-face interaction (63% in the student survey). This perception was related to challenges met in various areas, such as planning, coordination and coherence. Teachers and students both agreed that, for at least some competences, this less favorable learning experience could have been more related to the sudden switch due to the crisis itself rather than the limitations associated with the online methods.

> “… *I think that, if things were conducted properly, with clear guidelines easily accessible by everybody, good organization and no last-minute changes, there should not be much problem and results would not have differed much from what is usual*.” (student)

Despite this perception, the scores obtained in the final exams following the online teaching experience have not changed significantly from those obtained in previous years, although this does not constitute a guarantee of learning quality [14,15].

### Teaching methodology

Students and faculty both missed the pre-pandemic face-to-face teaching model. This model combined various methodologies: lectures attended by the entire class of up to 90 students, case and problem-based learning, simulation and skill labs delivered in smaller groups, etc. Despite the efforts to port all these learning methods to their virtual counterparts, the transformation process generated, at least during the initial stages of deployment, confusion and lack of confidence regarding the expected learning outcomes.

The experience gained through the process highlights again the importance of the teaching methodology in stimulating active learning. It also underlines the need to rethink the time devoted to each of the learning activities and focus instead on the learning objectives. Such process involves reducing the time spent on lectures, as previously reported [16], and calls for the use of side activities that help consolidate learning.

As far as the class design is concerned, following an orderly sequence of contents, splitting lectures with other activities, allocating time for Q&A and making use of engaging activities allowed for immediate feedback between students and the teacher. In this regard, the activities best rated among students were those that took place live (synchronous), in line with what others have reported [17]. This was corroborated by the results of the survey, which rated synchronous learning and synchronous case-based methods with the highest scores (3.9/5 and 3.4/5, respectively).

> *“Synchronous [learning methods] made it easier for me to interact and address questions and clarifications” (professor)*
>
> *“*… *dealing with clinical cases [the use of synchronous learning methods] obviously encouraged you to ask more questions and [obtain] more feedback” (student)*

Faculty and students also stressed the importance of being able to keep eye contact in order to “not feel alone in front of a screen”, even if it was not constant. Surprisingly, some students described participating in online sessions more actively compared to face-to-face classes, suggesting that those with a more introvert type personality could feel more confident while participating in online classes, especially through chat.

> *“We have been told by many professors, at some point or another, that they want to see our faces” (student)*
>
> *“Following class from home alone, without the presence of fellow classmates and teachers, is demotivating. No one to discuss cases side by side with” (student)*

### Online resources

The use of online resources was generally well perceived as long as teachers showed enough skills in their use and could take advantage of their full potential. The diversity in the type of online resources used caused discomfort among some students. Therefore it is important to adequately train faculty and students in these “digital competences”, which may become useful in their subsequent process of self-learning.

Resources that facilitated participation and feed-back, particularly those that were delivered synchronously, were among the most welcomed. The same applied to audiovisual resources, such as short video clips showing specific cases or particular clinical aspects.

> *“The best thing for me has been Collaborate*^*™*^ *[delivered] live, because you can write anything on the screen later or while in class” (student)*
>
> *“A live [online] class, even better when recorded, is the closest thing to a real class, especially when the teacher encourages interaction. Also, when the class is*
>
> *recorded it makes it easier for me to manage my time and listen to the class whenever I want*.*” (student)*
>
> *“*… *you can run live surveys, such as “I agree / don’t agree”, the possibilities are endless*…*” (student)*

On the other hand, the forums, which had been available to faculty on the virtual platform for a long time, were discovered and put to use by some of them, increasing the communication and participation of students, despite the fact that these tools did not receive the highest scores.

> “. .. *also, the good thing about forums is that they allow the students to become teachers, that is* … *students themselves respond to the questions posed by their peers. Obviously, [this needs to be] monitored by a teacher*” *(faculty)*

Students and teachers also stressed the importance of having the proper conditions at home available in terms of space, technical equipment and family dynamics to facilitate online learning and teaching.

### Evaluation

Shortly after the confinement began, the DO asked faculty to review the evaluation criteria in their subjects increasing the number and weight of activities that involved a continuous assessment. This measure had two objectives in mind. On one side, it was intended to improve the level of follow-up and attention given to students. On the other, it was directed at anticipating the need for a fair method of assessment due the difficulties and limitations inherent to an online “final exam”, particularly in the event a final face-to-face evaluation could not be possible, as it turned out to be the case. From a teacher’s perspective, the need to implement these changes in such a short period of time became a challenge of its own, as it was executed with varying levels of uncertainty, a sense that was often transferred to students.

> *“*… *I have been asked to strengthen those activities involving continuous evaluation, which is something I did not have in mind* … *and this has added stress to the situation” (faculty)*
>
> *“They were assigning many tasks to us and we were taking them as they came in, but, in the end, many hardly counted towards the final grade*…*”(student)*
>
> *“It has become increasingly difficult for me to evaluate students. I have lost the evaluation component tailored to assess the individual progress; it is no longer there with this system” (faculty)*

### Time management

Both students and faculty expressed that online learning is a flexible modality that facilitates time management. This has been emphasized by older students. Nevertheless, because online learning requires efficient time management and not all students show equal skills in this regard, we have become aware that time management is a competence that students need to be trained for, particularly during the first years in school.

One aspect that has been highlighted by students is the decrease of dead time due to waits and commuting, which many admittingly not always took advantage of due to poor time management. Deficits in proper planning and miscommunication may have also contributed to this fact.

> *“It has helped me to learn to manage my own time” (student)*
>
> *“*… *has allowed me to study at my own pace and manage my time, although on occasion I ended up tangled in confusion” (student)*

### Workload

An additional consequence of the coordination deficit was the increased workload on students, which contributed to the aforementioned stress. Some faculty reacted to their perception of a lack of commitment on behalf of students by assigning them new tasks and projects, perhaps with a relatively small weight in the final grade. Students often considered this burden to be excessive, which was confirmed by the survey results (workload considered “high” or “very high” by 87.7% of respondents), placing this item among the worst rated (score 0.7/5).

> *“I have lived through university with an intensity that I had never experienced before, much due to the amount of workload [given to us]* … *It was of course very difficult at a personal level, given the situation we were going through. But I felt the university was all I could focus on” (student)*

At the same time, faculty also pointed out that the sudden transition to online teaching resulted in a greater workload than usual, on top of the high demands of their practice and family obligations. Some additionally expressed a lack of digital skills to carry out the switch, which resulted in choosing the simplest resources available, as has been reported in other studies [17-19].

### Student Motivation

Students reported difficulties in staying motivated (score: 2.3/5). They related this to the feeling of uncertainty, lack of coordination, excessive workload, as well as the absence of university social life and decreased relationship with fellow students and faculty, which led to a lower sense of belonging. Some of the faculty also claimed a “feeling of loneliness” due to the absence of direct contact.

Both students and faculty agreed that when teachers are passionate about what they are teaching and stay motivated despite the difficulties of the environment, that boosts student’s motivation as well.

> *“I’ve noticed that subjects that do really well are those where you would notice a teacher behind that stays highly motivated” (student)*

According to students, the most helpful teaching activities to stay motivated were those that facilitated involvement and student participation, such as case resolution, gamification and the use of questionnaires and surveys.

### Participation

The students participated in the decision-making process from the beginning of the crisis through their class representatives. Overall, however, they felt they were left out of many decisions.

On one hand, the need for urgent action required the use of participation channels that differed from those employed under regular circumstances. On the other, the crisis situation brought to test the level of endorsement of the class representatives, which was occasionally questioned by the student community at large due to miscommunication.

> *“*… *a meeting was held between faculty and students to reach an agreement* …*things have been running pretty smooth ever since…. (student)*

As far as participation regarded as student engagement in the different learning activities, this has been discussed in the sections that cover teaching methodology and online resources.

### Teacher-student relationship

Students and faculty were accustomed to receiving a standard of personalized advice and guidance considered by many as distinguishing features of the education provided by our University. Both have missedthe close teacher-student relationship in class and outside the classroom, as well as the social life on campus.

> *“The interaction with students is* very *different*, … *I have experienced it witha level of sadness, likea loss… it’s not the same*.*” (faculty)*
>
> *“Online training implies social isolation. it was required in this case, of course*,
>
> *but being able to contact face-to-face with your peers and teachers is a different ball game” (student)*

Consequently, teacher-student relationships scored low in the survey, although significantly worse in clinical courses compared to pre-clinicals (1.9/5 vs. 2.3/5, respectively). Such disparity could be attributed to differences in the profiles of teachers, students and teaching content between the two sections of the degree, aggravated by the health crisis [5].

### Final remarks

Interestingly, overall acceptance of distance learning among our medical students only scored fairly well, whereas other studies have reported better perceptions and attitudes from students towards this learning modality [20,21]. This difference could be explained by factors related to the specific educational and epidemiological context of emergency in which our study was conducted, including the abrupt shift from face-to-face to online learning, with little time to adjust to the new online training skills required, the involvement of many faculty in providing healthcare to COVID-19 patients, and the deep psychological and social impact of the pandemic among students, who experienced strict home confinement for more than three months.

In this regard, the results presented here may not be applicable to other contexts. The particular aspects of our cultural environment may have influenced the results obtained and should be taken into account when generalizing the conclusions to other cases. Another limitation affects the quantitative part of this study. Although the participation level for the survey was acceptable, as we collected responses from up to half of the number of students enrolled, the possibility of self-selection bias cannot be discarded.

Virtual teaching should preserve and enhance key aspects of learning such as student motivation and participation at all levels. This leads to the need to rethink some of the elements of teaching of a more organizational nature, such as course planning, coordination, communication and pedagogical coherence. In turn, the rethinking process should extend to other aspects identified in this study such as learning outcomes, evaluation, teacher-student relationship, time management and workload.

In this new scenario, teaching methodology and online resources are aspects of particular relevance in the learning process, and should be delivered synchronously and to their maximum potential with the proper training of both students and faculty.

## Data Availability

The datasets used and/or analyzed during the present study are available from the corresponding author on reasonable request.

## Acknowledgements

We would like to thank the medical students and faculty members at UIC School of Medicine and Health Sciences that participated in the focus and nominal group discussions respectively, as well as the medical students that responded to the survey.

## Author contributions

Conceptualization and methodology, MV, ME, MG, AB; qualitative and quantitative data collection, MV, ME; quantitative data analysis, PB; mixed methods analysis, MV, ME, MG; writing-original draft preparation, MV, ME, MG, PB, PM; review and approval of final draft, MV, ME, MG, PB, PM, AB.

## Ethics approval and consent to participate

The study was approved by the Ethics and Research Committee of the study site and written informed consents were obtained from all participants prior to initiation.

## References

1. WHO. Public Health Emergency of International Concern declared. Available from: https://www.who.int/emergencies/diseases/novel-coronavirus-2019/events-as-they-happen [Accessed September 3, 2020].

2. CCAES Centro de Coordinación de Alertas y Emergencias Sanitarias. [Update n. 219. Coronavirus Disease (COVID-19). 01.10.2020]. Available from: https://www.mscbs.gob.es/en/profesionales/saludPublica/ccayes/alertasActual/nCov-China/documentos/Actualizacion_219_COVID-19.pdf [Accessed October 20, 2020].

3. Ferrel MN, Ryan JJ. The impact of COVID-19 on medical education. Cureus. 2020;12: e7492.

4. Rolak S, Keefe AM, Davidson EL, Aryal P, Parajuli S. Impacts and challenges of United States medical students during the COVID-19 pandemic. World J Clin Cases. 2020;8(15): 3136–3141.

5. Rose S. Medical student education in the time of COVID-19. JAMA. 2020;323(21):2131–2132.

6. Dedeilia A, Sotiropoulos MG, Hanrahan JG, Janga D, Dedeilias P, Sideris M. Medical and Surgical Education Challenges and Innovations in the COVID-19 Era: A Systematic Review. In vivo. 2020;34(3Suppl):1603–1611.

7. Creswell JW, Plano Clark VL. Designing and Conducting Mixed Methods Research. Thousand Oaks, California: Sage Publications;2011.

8. Creswell JW. A concise introduction to mixed methods research. Thousand Oaks, California: SAGE publications;2014.

9. Brotons P, Virumbrales M, Elorduy M, Mezquita P, Graell M, Balaguer A. [Learning Medicine at distance?: perception of students confined by the COVID-19 pandemic]. Rev Med Chile. 2020. Accepted for publication. Spanish.

10. Bisquerra R. [Methodolody of research in education]. Madrid: Editorial La Muralla; 2004.

11. O’Doherty D, Dromey M, Lougheed J, Hannigan A, Last J, McGrath D (2018). Barriers and solutions to online learning in medical education - an integrative review. BMC Med Educ. 2018;18(1):130.

12. Dyrbye LN, West CP, Satele D, Boone S, Tan L, Sloan J, Shanafelt TD. Burnout among U.S. medical students, residents, and early career physicians relative to the general U.S. population. Acad Med. 2014;89(3):443–451.

13. Dyrbye L, Shanafelt T. A narrative review on burnout experienced by medical students and residents. Med Educ. 2016;50(1):132–149.

14. Chumley-Jones HS, Dobbie A, Alford CL. Web-based learning: Sound educational method or hype? A review of the evaluation literature. Acad Med. 2002;77(Suppl 10):86–93.

15. Regmi K, Jones L. A systematic review of the factors - enablers and barriers - affecting e-learning in health sciences education. BMC Med Educ. 2020;20(1):91.

16. Daud A, Bagria A, Shah K, Puryer J. Should Undergraduate Lectures be Compulsory? The Views of Dental and Medical Students from a UK University. Dent J (Basel). 2017;5(2).

17. Khalil R, Mansour AE, Fadda WA, Almisnid K, Aldamegh M, Al-Nafeesah A, et al. The sudden transition to synchronized online learning during the COVID-19 pandemic in Saudi Arabia: a qualitative study exploring medical students’ perspectives. BMC Med Educ. 2020;20(1):285.

18. Bączek M, Zagańczyk-Bączek M, Szpringer M, Jaroszyński A, Wożakowska-Kapłon. Students’ perception of online learning during the COVID-19 pandemic: a survey study of Polish medical students. Res Sq [Preprint] 2020. Available from: https://www.researchsquare.com/article/rs-41178/v1 [Accessed October 20, 2020].

19. Shahrvini BB, Baxter, Coffey CS, MacDonald BBV, Lander SL. Pre-Clinical Remote Undergraduate Medical Education During the COVID-19 Pandemic: A Survey Study. Res Sq [Preprint] 2020. Available from: https://www.researchsquare.com/article/rs-33870/v1 [Accessed October 20, 2020].

20. Peng Y, Pei C, Zheng Y, Wang J, Zhang K, Zheng Z, et al. A cross-sectional survey of knowledge, attitude and practice associated with COVID-19 among undergraduate students in China. BMC Public Health. 2020;20(1):1292.

21. Sandhaus Y, Kushnir T, Ashkenazi S. Electronic Distance Learning of Pre-clinical Studies During the COVID-19 Pandemic: A Preliminary Study of Medical Student Responses and Potential Future Impact. Isr Med Assoc J. 2020;8(22):423–427.

